# Therapist perspectives on the clinical utility of hand performance information from at-home egocentric video in outpatient neurorehabilitation: A multi-methods evaluation study

**DOI:** 10.1101/2024.09.27.24314512

**Authors:** Adesh Kadambi, Damian M. Manzone, José Zariffa

**Author notes:** **Corresponding author(s):** Adesh Kadambi, BEng, José Zariffa, PhD, PEng.

## Abstract

**Background:** Restoring hand function is a primary focus of rehabilitation after neurological injuries, such as stroke and spinal cord injury. However, monitoring hand use outside the clinic remains challenging. This study aims to evaluate how therapists perceive and would utilise information from a clinical decision support system (CDSS) that uses egocentric video to monitor patients’ hand use at home.

**Methods:** Five patient-therapist dyads were recruited. Patients recorded daily activities using head-mounted cameras. Therapists reviewed dashboards of processed video data from their patients and completed semi-structured interviews and structured questionnaires. A multi-methods approach with thematic analysis was used to evaluate the CDSS’s clinical usefulness.

**Results:** Qualitative analyses revealed four main themes: therapists strongly preferred observational video data over quantitative metrics, valued insights into home environments that cannot be captured in-clinic, identified ways the information could tailor therapy to real-world activities, but noted practical implementation challenges including time constraints and patient burden. The system achieved high usability scores (mean SUS: 80.0, 85-89th percentile).

**Conclusions:** This study demonstrates the potential of using egocentric video to inform clinical decision-making in neurorehabilitation, particularly for hand function. The strong preference for video over metrics suggests clinical decision support systems should prioritize interpretable, observation-based information aligned with clinical reasoning processes. Despite implementation challenges, therapists across technical familiarity levels expressed trust in the system and willingness to use it regularly. These findings indicate that egocentric video systems can bridge the clinic-home divide when designed to match interest-holder priorities.

## Introduction

Neurological injuries and conditions commonly result in substantial loss of upper limb function, which greatly limits an individual’s ability to perform activities of daily living (ADLs). Across diverse neurological populations, impaired hand function consistently ranks as a primary rehabilitation concern. For instance, individuals with spinal cord injury (SCI) consistently report regaining hand and arm function as the top recovery priority [1]. Similarly, for individuals who have experienced a stroke, data suggest upper-limb strength is a major predictor of quality of life [2]. Therefore, restoring hand function is a primary focus of rehabilitation efforts, as it directly impacts the ability to regain independence and quality of life.

Regular and accurate monitoring of hand function is essential for promoting recovery [3]. It enables clinical assessment and progress tracking, thereby allowing therapists to adjust treatment plans, and ensure therapy addresses real-world functional needs [3]. However, monitoring this function outside of the clinic remains a significant challenge [4, 5]. Conventional outpatient therapy is typically limited to face-to-face assessments and patient self-reports, which often fail to capture the complexity and variability of real-life hand use at home [6–9]. Patient self-reporting also relies on an individual’s ability to recall activities performed since their last therapy session and is subject to reporting biases, such as cognitive deficits and social desirability [10–13]. These limitations prevent clinicians in outpatient settings from gaining a comprehensive understanding of how patients perform ADLs in their everyday environments, thus hindering the ability to tailor therapy plans on the basis of real-world functional performance.

Wearable technology offers an avenue for measuring hand function outside of the clinic. In particular, head-mounted egocentric cameras can provide rich, contextual insights into patients’ hand function during ADLs [14–16]. However, clinicians have shown reluctance to adopting new technologies in their practices, citing concerns about time constraints and uncertainty regarding clinical value [5, 17–19]—factors identified by the Technology Acceptance Model [20] as primary barriers to adoption [21]. Digital tools, such as clinical decision support systems (CDSS), have demonstrated success in implementing evidence-based guidelines at the point of care and improving patient outcomes in various healthcare domains [22], including rehabilitation [23, 24], chronic disease management [25], and prescription and medication safety [26]. However, this success hinges on whether they present information that is useful, interpretable, and actionable for clinicians [27–30]. Therefore, understanding how clinicians perceive and value specific types of information from wearable devices is crucial for developing systems that address time and value concerns that have previously hindered the adoption of new technologies in clinical practice.

We recently co-designed a CDSS dashboard with clinicians to present information about patient hand use captured through egocentric video recordings in multiple formats, including quantitative metrics and video footage of functional activities [31]. Our current study aims to understand how this information from egocentric videos, presented through the dashboard interface [31], is perceived by therapists in the context of their clinical practice and decision making. Specifically, we sought to evaluate whether therapists find the information useful for better understanding patient hand performance at home and whether it could inform clinical decision-making. These aspects have not been explored in prior work with egocentric video in rehabilitation, which has primarily revolved around developing research tools to measure outcomes in clinical studies [15, 16, 32] and exploring patient perspectives [33, 34]. Due to the need for us to investigate how this data might be useful to therapists in clinical practice, we evaluated this system with therapists treating patients with stroke and spinal cord injury as representative examples of acquired neurological conditions affecting upper limb function. Through this work, we seek to bridge the gap between digital health tool development and practical clinical application, ensuring that innovations can meaningfully enhance patient outcomes in neurorehabilitation settings.

## Methods

This study was approved by the Research Ethics Board of the University Health Network (UHN; Study #21-5019) and conducted in accordance with the Tri-Council Policy Statement. We employed a multi-methods design to evaluate the clinical usefulness of the information presented to therapists [35]. Qualitative data from semi-structured interviews were analysed using a thematic analysis to explore therapist perspectives in depth. Quantitative data from structured questionnaires and the System Usability Scale (SUS) were analysed using descriptive statistics. These two data sources were analysed separately, with quantitative findings used to corroborate and contextualize qualitative themes. This approach allowed for a comprehensive understanding of the system’s clinical utility and usability.

### Participants

We recruited 5 patient-therapist dyads from the Toronto Rehabilitation Institute at UHN using a snowball sampling approach. This sample size was deemed appropriate for this study due to its specific research question and a single analytically relevant participant category (i.e., therapists assessing clinical utility) [36]. Therapist-participants were eligible if they had expertise in upper limb rehabilitation and were actively treating patients who were either adult stroke survivors or adults living with SCI. Patient-participants were eligible if they were over 18 years of age, actively receiving therapy at UHN from the therapist-participant, and had self-reported impairment of hand function from SCI or stroke while retaining the ability to use their hands in some capacity for at-home ADLs.

The therapist group included 4 occupational therapists and 1 physical therapist with 13.8 ± 5.5 years of clinical experience (range: 8-20 years). All therapists were female. The patient group consisted of 2 stroke survivors and 3 individuals living with SCI. This mix of stroke and SCI patients allowed us to explore the system’s applicability across different types of upper limb impairments. All participants were assigned an alphanumeric code to anonymize transcript data and any excerpts in this article.

### Data Collection

Our data collection process followed a three-stage pipeline consisting of patient video recordings, dashboard generation, and therapist interviews, as illustrated in Figure 1.

**Figure 1:**
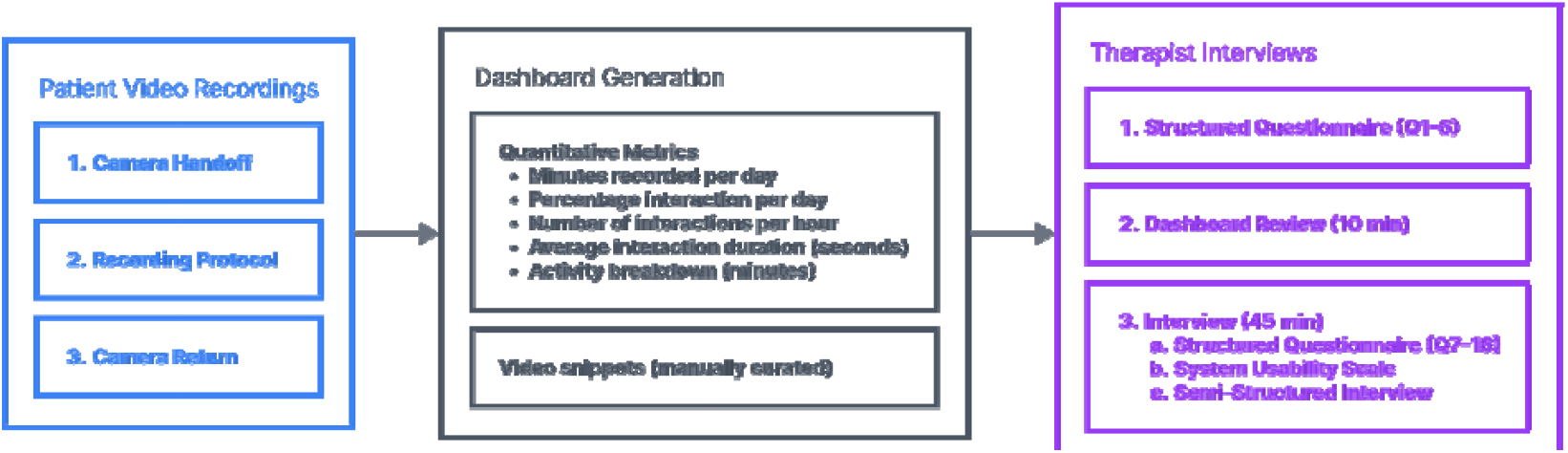
Overview of the data collection pipeline. Patient-participants recorded videos at home using a head-mounted egocentric camera. A dashboard was generated from the recordings, which included both quantitative metrics and manually curated video snippets. Therapist-participants reviewed the dashboard generated for their patient for approximately 10 minutes before proceeding with the interview.

### Video Recordings

Patient-participants were provided with head-mounted egocentric cameras (GoPro Hero 5, GoPro, Inc., San Mateo, CA, USA) to record their ADLs at home. The cameras recorded videos in .mp4 format at 1080p resolution and 30 frames per second. Patient-participants were instructed to record 1-hour sessions between 2-4 times per week for a minimum of 2 weeks. Following a previously reported protocol, recording times were agreed on by the investigator and participant in an initial interview [37]. The recording periods were selected to capture times when patient-participants performed tasks that were informative about their hand function, part of their regular routine, and did not raise privacy concerns. Examples included eating, food preparation, household tasks (e.g., laundry, putting away groceries, writing), or non-private grooming tasks (e.g., brushing teeth, combing hair). Participants had full control to start and stop recordings and could edit videos before returning them to the research team. The patient-participants recorded an average of 231.85 ± 75.49 minutes of video. Once the recordings were completed, the patient-participants returned the cameras to the researchers.

### Dashboard Generation

The recorded videos were processed to generate a dashboard [31] that provided therapists with information about their patient’s hand use at home. The complete dashboard generation and video processing pipeline is illustrated in Figure 2. Raw videos were segmented into 1-minute clips and sampled at 2 frames per second. We applied existing computer vision models to detect objects [38] and identify hand-object interactions (HOI) [39]. These detections were used to compute quantitative metrics and classify activities. *Minutes recorded per day* was computed using the metadata from the videos. HOI detections were used to compute *percentage interaction* per day (i.e., proportion of frames containing detected interactions), *number of interactions* per hour (i.e., distinct hand-object engagement events, defined as consecutive frames with detected interactions), and *average interaction duration* (i.e., temporal length of interaction events in seconds) [16]. The *activity breakdown* was generated using an ADL classification model [40] to categorize videos into seven ADL categories, and then manually verified for accuracy. *Video snippets* were manually curated from the 1-minute video segments by AK, with an intentional focus on tasks the patient struggled to complete or performed with compensatory strategies, as these were deemed most clinically relevant for informing therapy planning through prior conversations with therapists.

**Figure 2:**
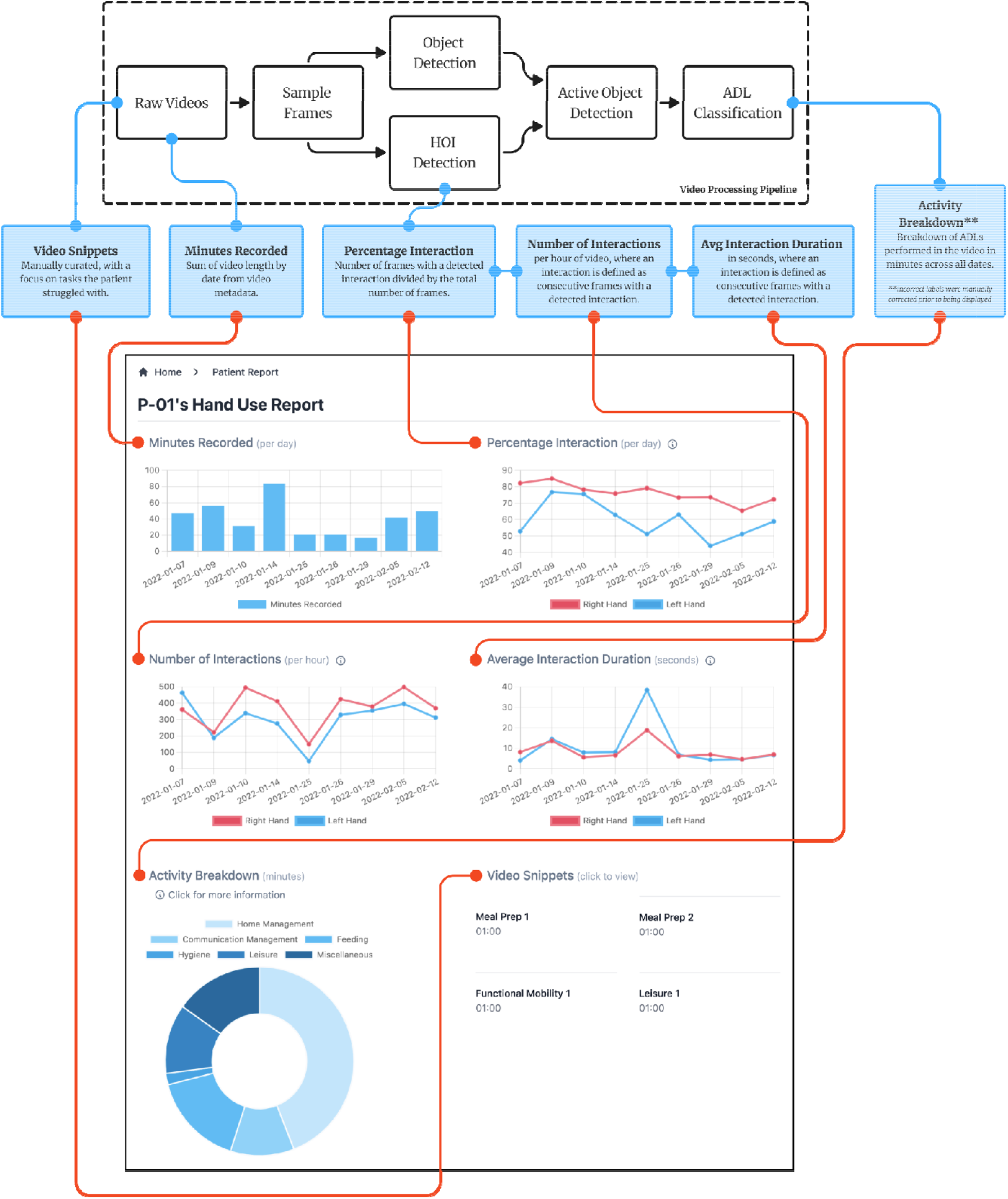
Dashboard generation pipeline. Raw egocentric videos were segmented into 1-minute clips and sampled at 2 frames per second. Object detection and HOI models identified active hand-object interactions. These features were used to automatically classify ADLs [40]. A sample dashboard for a patient-participant with synthetic data is displayed below the processing pipeline. Blue boxes show dashboard metrics and their derivation points. The red lines point to the specific user interface elements for each metric.

### Therapist Interviews

The final stage of data collection involved therapist-participants reviewing and providing feedback on the CDSS. This process began with a 10-minute review period where the therapist could familiarise themselves with the dashboard and review the information provided for their patient. This was followed by a 45-minute interview session consisting of structured questionnaires and a semi-structured interview. The structured questionnaires included 5-point Likert scale questions to assess technological affinity and perceptions of clinical utility and actionability, as well as a SUS questionnaire [41] to measure the usability of the CDSS. The SUS is a widely validated, reliable 10-item scale for evaluating system usability across diverse applications [42]. The semi-structured interview gathered in-depth qualitative feedback on the usefulness of the information and its influence on clinical decision-making. Four of the five patient-participants had completed their therapy block between one and three months prior to the therapist interview, while one remained in active therapy. This retrospective evaluation allowed therapists to reflect on how the information could have informed their clinical decision-making. All interviews were conducted by AK using an interview guide developed for this study and provided in the supplementary material.

### Data Analysis

We analysed qualitative and quantitative data separately and then presented findings together to provide a comprehensive understanding of the system’s clinical utility and usability. The data from the semi-structured interviews were transcribed verbatim and analysed using reflexive thematic analysis [43, 44]. Reflexive thematic analysis is an iterative, interpretive approach where themes are actively constructed by researchers through engagement with the data [44, 45]. This process involved two independent researchers (AK and DMM) coding the data to identify key themes and patterns related to the clinical usefulness and actionability of the information presented in the dashboard. Following initial coding, the researchers engaged in consensus coding, where they met to compare their findings, discuss any discrepancies, and reach agreement on the final set of themes and codes. JZ acted as an arbitrator for any unresolved discrepancies. This approach was employed to reduce individual bias and enhance the reliability and credibility of the analysis. The Likert scale questionnaire responses were analysed using descriptive statistics. The SUS scores were used to assess the usability of the dashboard format for presenting this information using the curved grading scale interpretation [46]. Quantitative findings were used to corroborate and contextualize the qualitative themes identified through thematic analysis.

## Results

### Thematic Analysis

Four main themes were constructed through our reflexive thematic analysis: (1) Data Interpretation Preferences, (2) Bridging the Clinic-Home Divide, (3) Tailoring Therapy Through Home Observations, and (4) Practical Realities of Implementation. Table 1 presents an overview of these themes and their subthemes.

**Table 1:** Overview of themes and subthemes for thematic analysis from semi-structured interviews.

| Theme | Subthemes |
| --- | --- |
| Data Interpretation Preferences | 1A. The Primacy of Functional Footage<br>1B. Navigating Novel Graph Presentations |
| Bridging the Clinic-Home Divide | 2A. Beyond Clinic Walls<br>2B. Goal-Oriented Data Collection |
| Tailoring Therapy Through Home Observations |  |
| Practical Realities of Implementation | 4A. Clinical Integration<br>4B. Another Burden? |

#### 1. Data Interpretation Preferences

This theme explores how therapists interacted with and valued the different types of data presented to them in the dashboard (i.e., graphs and videos). It highlights the distinction between more familiar, observation-based representations of data and novel data visualisation techniques.

##### 1A. The Primacy of Functional Footage

This subtheme captures therapists’ strong preference for video data and its ability to provide rich, functional insights into patients’ real-world activities, emphasising its alignment with how they already consume data they are presented with in the clinic (i.e., through patient observations). As one therapist highlighted:

> I would go right to the videos. The graphs were interesting in that I saw that he looked like he used his left hand more, but that probably wouldn’t give me that much information. I think the videos are the most telling, at least for me. Visual, more direct application for me. (T05, OT)

This sentiment was also echoed by others: “To be honest, I really liked the videos, because that, as much as the number tells you, it doesn’t actually tell you the quality and the pattern that she’s actually using her hand” (T01, PT).

Overall, while the graphical data could be a good overview, therapists believed the real value came from the videos: “The quantitative measures would give me an idea of how much he’s progressing in terms of quantity of affected hand use, […] and then just yeah the videos give me a lot of rich information on how I can adapt therapy” (T04, OT).

##### 1B. Navigating Novel Graph Presentations

In contrast, therapists expressed challenges interpreting the information presented in the graphs and found it difficult to integrate into their clinical reasoning or therapy planning in any way:

> Normally I don’t go through metrics or graphs or, this isn’t a typical format for me to obtain or analyse information being more like a frontline clinician. So just again, my eyes aren’t as familiar with this as from what my role has been in the past, like 20 years here. This isn’t a regular part of how we’re interpreting our patient information. […] I think there just needs to be more specification around [the graphs] or just more defining parameters just for it to have any value. […] like if we’re using this at [intake] and then discharge, I would want it to be as comparable as possible. (T02, OT)

#### 2. Bridging the Clinic-Home Divide

This theme explores the unique insights that therapists gained from the CDSS about patient home environments and daily activities beyond the clinical setting. It also highlights different data collection approaches to make the information more useful to therapists.

##### 2A. Beyond Clinic Walls

Therapists emphasised the value of seeing videos of their patients performing functional activities at home “because the clinic is limited as a simulation of what occurs at home” (T04, OT). And since they “cannot go to every patient’s home, in a way [these videos] bring the patient’s home to [them]” (T03, OT). As one therapist noted:

> Part of our OT assessment does include the physical environment, so we are asking about obviously the basic layout, and how you’re getting around, and how you’re accessing different rooms, but as far as, a lot of times the physical environment also does just limit the potential for how much they can do. So, yeah, that’s just an extra layer I would say. But as far as thinking about whatever recommendations we might make for home modifications, or even how he most safely accesses his space, it’s just that opportunity to see a patient, again, outside of a very sort of generic clinic space or setting. (T02, OT)

##### 2B. Goal-Oriented Data Collection

While patients were instructed to record typical daily activities following the protocol outlined in [37], therapists expressed a preference for focused data collection aligned with patients’ specific goals and priorities, while also being able to compare how a patient is able to perform different functional activities across time points to better track progress over time:

> It would be useful to me to be able to see the change from one time-point to the next when it’s… like if this was a pre-therapy video, then I would want to be able to compare it readily to mid-therapy and post-therapy video. And I guess it would be nice to have some alignment between what his functional goals are, because for instance one of his goals initially was to be able to like open a ring box so I would have him videotape at home like how he’s performing that goal, or his approximation of that goal, like opening a box at home. I would want to see how he’s progressing with things that are part of his goals. (T04, OT)

#### 3. Tailoring Therapy Through Home Observations

This theme discusses how the new insights gained from the CDSS could be used to adjust therapist recommendations and therapy plans based on observed real-world patient behaviour. One therapist mentioned the videos provide much greater detail than their initial assessments and could help with task specific practice:

> We have like tedious notes as far as our initial assessment, but we would still certainly not be able to get that level of like, “So, okay, when you go in your kitchen…”. This really gives, if we want to break down the tasks even more, it definitely would lend itself really nicely to helping with more specific practice therapy sessions, or breaking down the task and say, “Okay, now let’s work on your left hand reach and then reaching across your body with your right hand to open the microwave”, or just some of the details definitely could help with the problem solving or maybe modified devices. (T02, OT)

Another therapist described how some of the activities she observed in the videos could be a safety hazard and that they would modify exercises in clinic to prevent a fall in the future:

> I love it […] you open my horizon as a therapist. Wow, [patient name] is doing some… one big help your project is… would be helpful in falls prevention because this patient is taking risk. I know the left leg is not working, so why not move your walker closer and then you grasp… and if she had… so I’m gonna modify my therapy by looking at those and improve my treatment so your project definitely will help us to improve the results. (T03, OT)

#### 4. Practical Realities of Implementation

This theme explores the practical considerations and potential barriers to implementing the dashboard in clinical practice. It encompasses concerns about time constraints for clinicians and the additional burdens placed on patients, highlighting the complexities of integrating new systems into existing clinical processes.

##### 4A. Clinical Integration

Therapists expressed varied perspectives on how they could use this system in their current practice. One therapist viewed the tool as being akin to reviewing patient charts in the electronic health record (i.e., Epic) prior to a session:

> I’d prefer to first look at it when I’m not in therapy. I think it’s very quick to extract information from this, so I don’t think this takes me any longer than you know, logging into Epic and doing all these things I have to do to document the session. […] Yeah, I don’t think it’s too time-consuming. I think it’s very valuable information. (T04, OT)

While another believed they did not “have a ton of time for it” (T02, OT), but considered “[looking] at it with the patient and [reviewing] it during the session” (T02, OT). As a result, one therapist questioned how often they would even use it: “I don’t know how often I would do it. If it was available, if there was something you really wanted to know, how are you actually doing this? Then I could see that coming into play.” (T05, OT).

##### 4B. Another Burden?

Therapists also mentioned potential challenges for patients:

So for someone who doesn’t really have any support, and for someone who doesn’t have support and a low level arm, like there’s no way they’re gonna get a GoPro on their head and strap it properly and turn it on and make sure it’s on and off, and you’re not gonna get them to do that for four weeks. So I think it’s, it’s great if, if you pick the right person to do it with, if not, I don’t know how helpful it would be. (T01, PT)

#### Quantitative Findings

In addition to the qualitative interviews, therapists completed a questionnaire about technology affinity and the clinical usefulness of this system in their practice (Figure 3). The first six questions aimed to understand a therapist’s affinity towards technology, and were answered before the review session began, while the remaining questions targeted their thoughts on our CDSS after the review period concluded. Regardless of therapists having varying levels of technology use and affinity in their lives (Q1-6), they all found the CDSS to be a useful tool for understanding patient performance at home (Q12), would use it regularly (Q7), and indicated that it had the potential to influence therapy planning (Q15).

**Figure 3:**
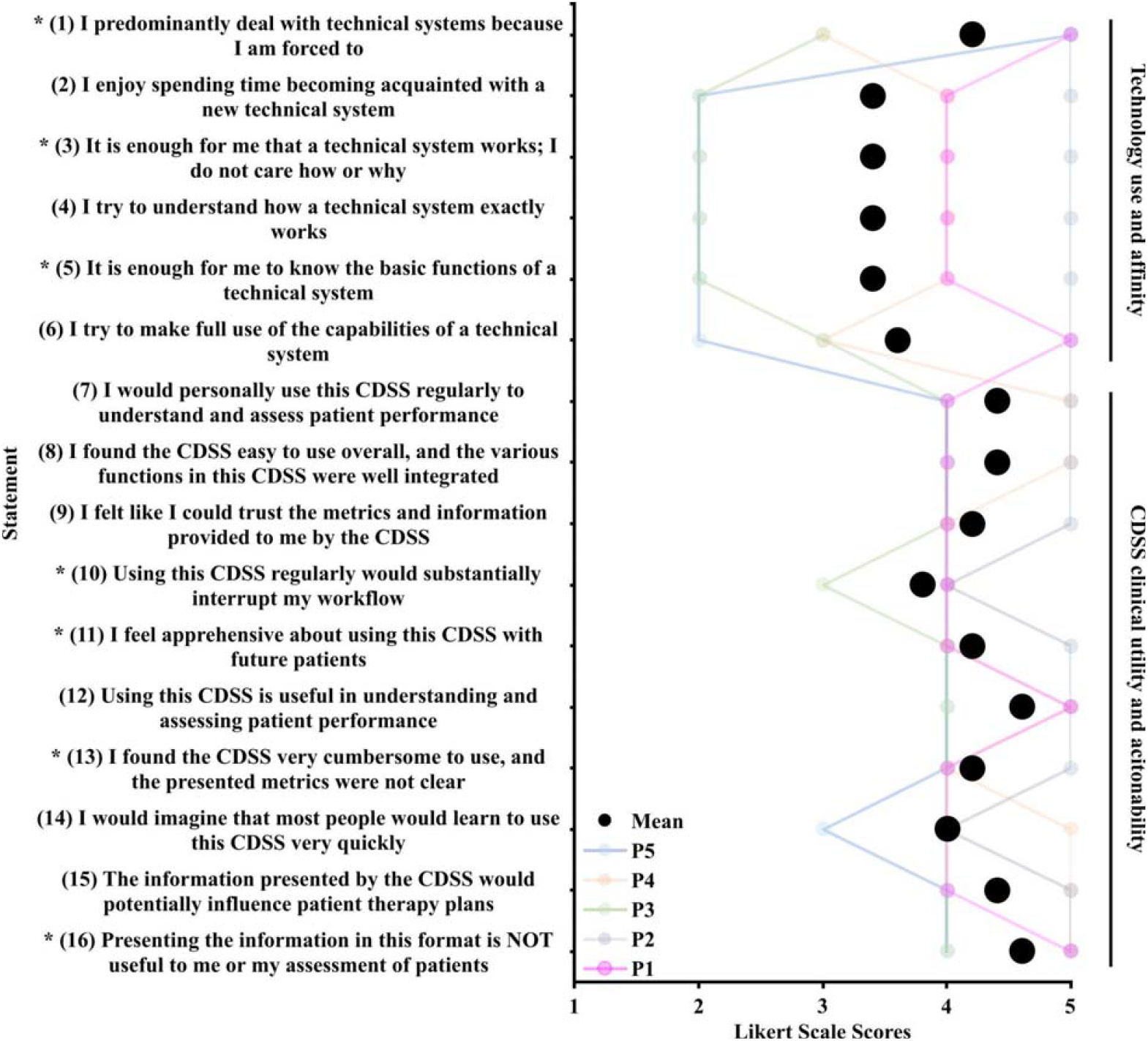
Therapist responses to the questionnaire regarding technology use and affinity (Q1-Q6) and clinical utility and actionability of the CDSS (Q7-Q16). Mean Likert scale scores are represented by black filled circles and the responses from each participant are represented by individually connected coloured lines. Likert score interpretations for the different questions are provided in Appendix. Statements with an asterisk (*) are reverse coded.

Our participants reported a mean SUS score of 80.0 (95% CI [65.95, 94.05]), placing our system in the 85-89th percentile range [46]. All individual participants scored above 68 (Figure 4), which represents the 50th percentile SUS score [46]. These results indicate above-average usability for our system overall.

**Figure 4:**
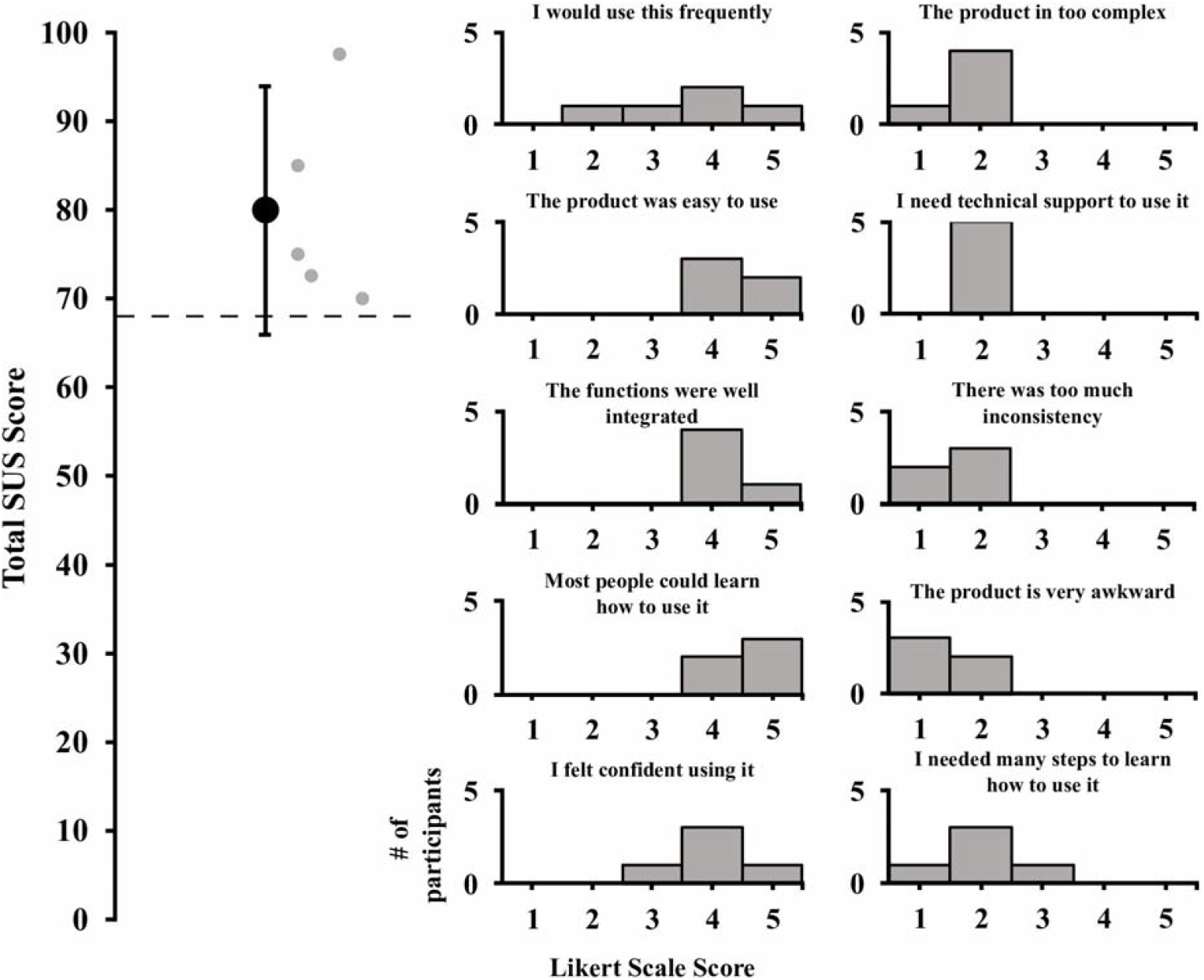
System Usability Scale (SUS) results. The left panel displays the total SUS scores. The mean score is represented by a black circle with error bars representing 95% CI. Individual participant scores are represented by the smaller grey circles. The dashed line is displayed at a total SUS score of 68, which is considered the average score at the 50th percentile [46]. The right panel displays the distribution of responses for all participants and for all statements. Note that the titles over each graph have been abbreviated for simplicity.

## Discussion

This study aimed to evaluate whether information about hand use in the home environment derived from egocentric video was perceived by therapists as being clinically useful in the context of outpatient upper limb neurorehabilitation. The information presented was well-received and gave therapists the ability to view patients performing functional activities and ADLs in their home environment, with the potential to result in more tailored therapy in the clinic. However, there were clear preferences for specific types of data and practical challenges for implementation.

Therapists consistently expressed a strong preference for video footage over graphical representations of data. The videos enabled therapists to observe real-world hand use and understand patients’ functional capabilities in their home environments, allowing them to adapt therapy based on direct observation of performance. For example, therapists reported the videos helped them break down tasks into specific movements performed at home, or provide more specific recommendations based on safety concerns they observed in the videos. This preference is supported by the high Likert scores related to the dashboard’s usefulness in assessing patient performance (Q7, Q12). This aligns with existing literature that underscores the importance of direct observation in clinical decision-making, where functional video footage provides critical context, such as movement quality, that is often lost in abstract numerical data [47, 48].

Conversely, several therapists found the graphical data more challenging to interpret and reported that the metrics were unclear and less directly applicable to their clinical reasoning. Concerns were raised about how quickly someone could learn how to use this information (Q14), suggesting a lack of clarity in how the data were presented, and its potential to disrupt workflow (Q10). This preference for video over graphical data likely reflects both the familiarity of video-based observation in clinical practice and the need for more intuitive data visualization design.

Our prior work [16] had focused on developing novel outcome measures for clinical research and demonstrated that quantitative metrics like percentage interaction and number of interactions were correlated to higher Upper Extremity Measurement Scale (UEMS) [49], Spinal Cord Independence Measure III (SCIM) [50], and Graded Redefined Assessment of Strength, Sensibility and Prehension (GRASSP) [51] scores. However, therapists in the current study found these very metrics less clinically actionable than raw video footage. This finding highlights the different considerations involved in using novel assessments as tools for clinical research or for direct care provision. For the therapist-participants in this study, the most useful aspect of this system was its ability to bridge the gap between the clinical and home environments. Therapists unanimously agreed that the clinic could never fully simulate the complexity of patients’ home environments and while they “cannot go to every patient’s home, in a way [these videos] bring the patient’s home to [them]” (T03, OT). Rather than presenting more sophisticated metrics, future systems may need to facilitate efficient navigation and interpretation of video content itself to better align with clinicians’ existing workflows and mental models.

An important consideration in developing home monitoring systems is ensuring alignment between patient and clinician perspectives. Our prior work has systematically explored these perspectives, first investigating patient acceptability and preferences [33, 34], and now examining therapist perspectives on clinical utility. The perspectives of patients from these prior studies and therapists from our current study converged on recognizing the fundamental value of home observation, indicating that clinic settings cannot fully capture home function, thereby supporting the core premise of home monitoring. Videos of patients performing functional activities at home gave therapists crucial insights into the physical and environmental barriers that patients encounter while performing real activities, which might not be evident in the controlled setting of the clinic. This ability to remotely observe home environments offers a promising alternative to traditional home visits, which are often limited by time and resource constraints even though they result in better patient outcomes [52–55]. The high score for statements indicating trust in the information (Q9) and the belief that it would influence future therapy plans (Q15) demonstrates the overall utility of our system in clinical settings.

However, despite recognizing the clinical value of the information provided by this system, therapists also highlighted several practical challenges related to its implementation such as clinical time constraints and patient burden. While some therapists felt the dashboard was easy to integrate into their existing chart review time, others worried that reviewing the data would add too much to their already busy schedules (Q10). There were also concerns about the burden on patients with limited physical or technological support, which aligns with previous findings, where individuals living with SCI and stroke expressed preferences for cameras that were less noticeable and lighter to minimise self-consciousness and discomfort [33, 34]. Some therapists also noted that certain patients might struggle to use the necessary equipment to capture their daily activities. These concerns echo the existing broader implementation science literature on barriers to adoption of mobile health and remote patient monitoring devices in healthcare, with disruption to clinician workflows and patient training and/or willingness to use these technologies playing the largest role in adoption rates [56–59]. Therefore, future iterations will explore ways to automatically highlight interesting video snippets for therapists to view. Additionally, future work should aim to design egocentric cameras that are easier to use by individuals with hand impairments and increase their willingness to use and record (i.e., lighter, less bulky, etc.).

## Limitations

While this study provides valuable insights into the clinical potential of the information presented by the CDSS, there are several limitations to consider. First, the small sample size of only five clinicians from a single hospital network limits the generalizability of our findings. This sample size was partly constrained by the number of eligible participants at our institution and the complexity of recruiting patient-therapist dyads, which is more challenging than recruiting therapists alone. Despite this limitation, the choice of 5 dyads was reasonable given that this study has a specific research question and a single analytically relevant participant ‘category’ (i.e., therapists) [36]. All participating therapists were female, and while we acknowledge that sex/gender may have influenced the findings of this study, our sample reflects the female-dominant nature of the field [60, 61]. Detailed patient demographics (e.g., age, time since injury, level of impairment) were also not collected, as this study focused on therapist perspectives regarding the clinical utility of the information system, evaluated in a manner agnostic to patient-specific characteristics.

Additionally, there is a potential for selection bias, as therapists more inclined to engage with and adopt new technologies may have been more likely to participate. While the quantitative questionnaire showed a wide range of responses in the technology affinity section (Q1-6), suggesting a variety of comfort levels with technology, it’s noteworthy that even therapists with lower technology affinity provided positive feedback on the CDSS. This indicates that the CDSS’s potential value was recognized across different levels of technological comfort. However, it’s important to acknowledge that the overall positive reception may not fully capture the perspectives of therapists who are generally less comfortable with technology and chose not to participate in the study.

Finally, although the CDSS was well-received in interviews, therapists did not have extended opportunities to integrate it into their daily practice. The video snippets were also intentionally curated to focus on tasks patients struggled with. This approach may have biased the presentation toward deficits rather than abilities, though therapists could also view the quantitative metrics which captured all hand use regardless of difficulty. This limited exposure means that the feedback gathered may not fully capture the tool’s long-term usability or the practical challenges it might pose once embedded in everyday workflows.

## Conclusion

This study demonstrates the potential of a system that uses egocentric video to provide therapists with home-based hand use data about their patients to inform clinical decision-making in a neurorehabilitation setting. While therapists found the information useful—particularly video data for assessing patient performance in a naturalistic setting—challenges related to integrating the system into clinical workflows were evident. The primary concerns centred around the clarity of graphical data and the perceived time burden that regular use might impose on therapists. Despite these hurdles, therapists across varying levels of technical familiarity expressed trust in the information provided by the system, indicated that they would use it regularly to assess patient performance, and believed it could enhance their ability to tailor therapy plans to real-world patient activities.

Reducing time demands through features like interpretations of graphical data or providing more emphasis on video data may help mitigate the barriers identified. Ultimately, the adoption of wearable egocentric cameras could facilitate a more seamless connection between face-to-face assessments and real-world functionality, contributing to more effective and personalised rehabilitation. However, further research is necessary to explore the long-term impact of this system in diverse clinical environments and its role in broader healthcare settings.

## Data Availability

All data produced in the present work are contained in the manuscript.

## List of abbreviations

SCI: Spinal Cord Injury
ADLs: Activities of Daily Living
CDSS: Clinical Decision Support System
UHN: University Health Network
SUS: System Usability Scale
OT: Occupational Therapist
PT: Physical Therapist

## Declarations

### Ethics approval and consent to participate

This study was approved by the Research Ethics Board of the University Health Network (Study #21-5019) and conducted in accordance with the Tri-Council Policy Statement: Ethical Conduct for Research Involving Humans (TCPS2). Informed consent was obtained from all individual participants included in the study.

### Consent for publication

Not applicable

### Availability of data and materials

The datasets generated and/or analysed during the current study are not publicly available due to privacy concerns regarding identifying information in home-recorded videos and interview transcripts but are available from the corresponding author on reasonable request.

### Authors’ contributions

AK recruited all participants, conducted and transcribed all of the interviews, performed data analysis, and drafted the manuscript. DMM contributed to data analysis, created figures, and revised the manuscript. JZ contributed to study design and data interpretation, and revised the manuscript. All authors read and approved the final manuscript.

## Acknowledgements

Not applicable

## Clinical Trial Number

Not applicable

